# Complexity of Coronary Disease and a Genetic Risk Profile in Patients with Type 2 Diabetes in Colombia

**DOI:** 10.1101/2024.12.02.24318358

**Authors:** German Camilo Giraldo-Gonzalez, Jheyson Jair Fuentes, Juan Carlos Sepúlveda-Arias, Carlos Eduardo Castaño

**Affiliations:** Health science doctorate, universidad de Caldas; Pureza de corazón unidad cardiología. Universidad de Antioquia; Grupo Infección e Inmunidad, Facultad de Ciencias de la Salud, Universidad Tecnológica de Pereira, Pereira, Colombia; Facultad de Ciencias de La Salud, Universidad de Caldas Manizales, Colombia

**Keywords:** Diabetes mellitus, diabetes complications, human genetics, coronary disease, cardiovascular disease, polymorphism, single nucleotide

## Abstract

**Background:** Coronary artery disease (CAD) remains a leading cause of mortality in patients with type 2 diabetes (T2DM). Both conditions share genetic and environmental etiologies, making T2DM a unique risk factor for CAD. Despite advancements, no highly accurate genetic or clinical prediction models exist to identify patients at high risk for complex coronary disease.

**Methods:** A cross-sectional genetic epidemiology study was conducted at a cardiovascular center in Manizales, Colombia, involving 106 T2DM patients with documented coronary heart disease (CHD). Genetic analysis focused on 11 single nucleotide polymorphisms (SNPs) using polymerase chain reaction (PCR) and TaqMan assays. Clinical and genetic data were analyzed to assess associations with CAD complexity, defined as monovascular (one vessel) or polyvascular (≥2 vessels) involvement. Statistical methods included bivariate analyses, binary logistic regression, and Bonferroni correction for significance.

**Results:** Among participants (mean age 68.1 ± 8.3 years; 53.7% male), 33 had monovascular disease, and 73 had polyvascular disease. Male sex (p = 0.016) and a significant genetic profile (PRP-5: rs1412830-CDKN2A/B, rs2074192-ACE2, rs4420638-APOE, rs646776-CELSR2, rs7903146-TCF7L2) were associated with polyvascular disease. PRP-5 demonstrated an odds ratio (OR) of 5.2 (p = 0.007) for complex CAD. Other metabolic and clinical parameters showed no significant differences between genetic risk groups.

**Conclusions:** This study identifies a polygenic risk profile (PRP-5) associated with complex CAD in T2DM patients, independent of traditional clinical factors. These findings support the hypothesis of genetic contributions to CAD severity and suggest the need for longitudinal studies to evaluate this risk profile’s clinical utility. As the first cohort-based genetic study in Hispanoamerica, it highlights the high prevalence of relevant alleles in this population and provides a foundation for future research into personalized cardiovascular risk stratification.

**Subject Áreas:** genetics, cardiometabolic disease, Type 2 diabetes, coronary disease, cardiovascular diseases, polymorphism, single nucleotide.

## Introduction

Understanding the genetics of atherosclerosis and coronary artery disease (CAD) in patients with type 2 diabetes (T2DM) remains a hot topic. At present, biology-based approaches to diagnosing and managing cardiovascular disorders are needed. (1–3)Coronary artery disease continues to be a major issue worldwide, with no definitive way yet to significantly reduce its burden on patients, physicians, and healthcare systems. (4,5)

CAD remains the leading cause of death in individuals with type 2 diabetes. Both conditions are complex and have both genetic and environmental causes, making T2D a unique risk factor for CAD. Although several studies have explored the shared genetic pathways between T2D and CAD, the findings have been limited.(6–8)

The Human Genome Study Association, the UK Biobank,(9) among others, have identified multiple single nucleotide polymorphisms with potential associations between coronary artery disease and type 2 diabetes. (10–14) However, it is necessary to verify these results in smaller cohorts(15). It has been documented that patients with T2DM not only have a higher risk of CAD but also of more severe and complex coronary disease.(16,17) Currently, there is no highly accurate clinical or genetic prediction model to help us detect early which patients are at higher risk for CHD and its complexity.(2,18,19) Figure 1.

**Figure 1.**
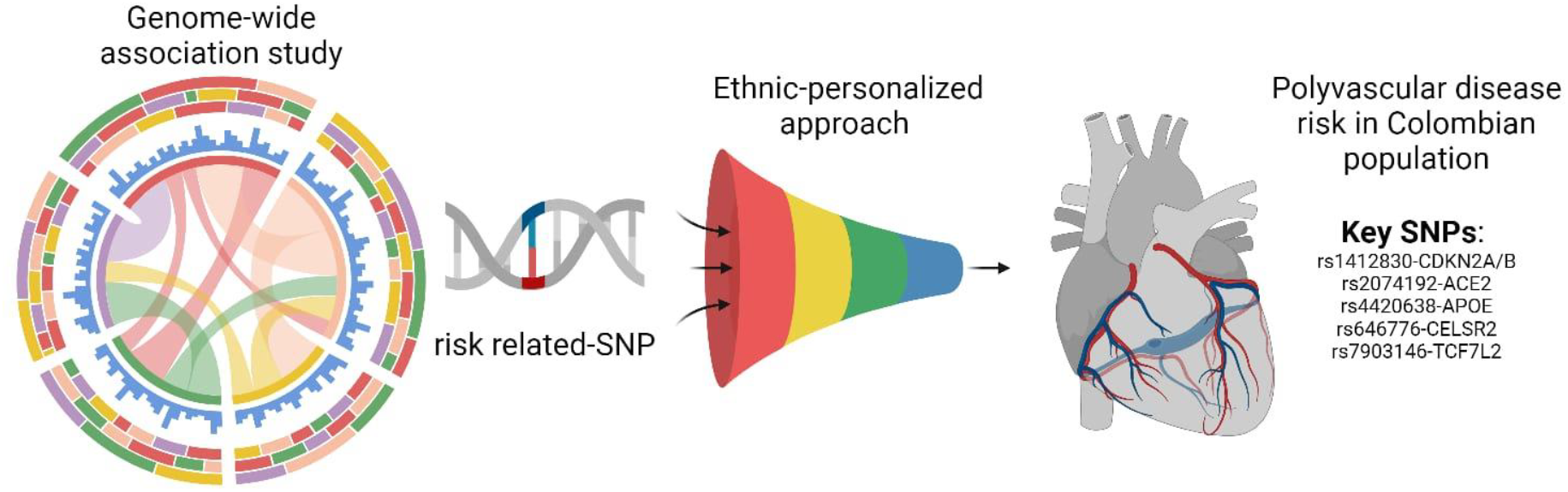
From genome-wide and big cohorts to personalized approach in complex coronary disease and validation in smaller cohorts.

## Methods

A cross-sectional genetic epidemiology study was designed, including all patients attending cardiology consultations at a specialized cardiovascular center in Manizales, Colombia, over three months. To be included in the study, patients had to be diagnosed with T2DM (according to the American Diabetes Association criteria) and coronary heart disease at some point in their lives.

Patients were excluded from the study if there was no documentation of invasive coronary angiography in their medical records. Once patients signed the informed consent, all registered information from the medical history was collected, and a sample was obtained via capillary puncture on Wassermann paper. Once the samples were collected, they were taken to the genetics laboratory for polymerase chain reaction (PCR) analysis using a TaqMan Assay kit.

### Statistical Analysis

Prevalence analyses were performed for the SNPs, according to their nature (homozygous or heterozygous), as well as the prevalence of clinical risk factors. Continuous variables were tested for normal distribution and are presented as mean ± standard deviation, while categorical variables are presented as percentages. Bivariate analyses were conducted using Chi-square (X^2^) goodness-of-fit tests as appropriate. Data collection was performed using a pre-designed tool with SPSS. Binary logistic regression was used to calculate odds ratios (ORs) for clinical risk factors and polymorphism reports. Statistical significance was set at a p-value of less than 0.05, and Bonferroni correction was applied.

For the association between clinical characteristics and risk genotype, patients were classified according to the complexity of coronary disease: monovascular (involvement of a single vessel) and polyvascular (involvement of two or more vessels). A frequency model was used to evaluate whether one, several, or any particular group of the 11 risk alleles might be related to more complex coronary disease.

## Results

A total of 106 patients were included in the analysis. The average age was 68.1 ± 8.3 years, and 53.7% were men, who also had more complex coronary artery disease than women (p 0.016). When evaluating the complexity of coronary artery disease, 33 patients had monovascular disease, and 73 patients had polyvascular disease. Regarding clinical factors, 75.4% of the patients had the three major risk factors (hypertension, T2DM, and dyslipidemia), 92% had hypertension, 80% had a history of smoking, 29.2% had a family history of premature cardiovascular disease, and 23% had chronic kidney disease. Table 1 shows the participants’ characteristics according to the complexity of coronary artery disease.

**Table 1.**
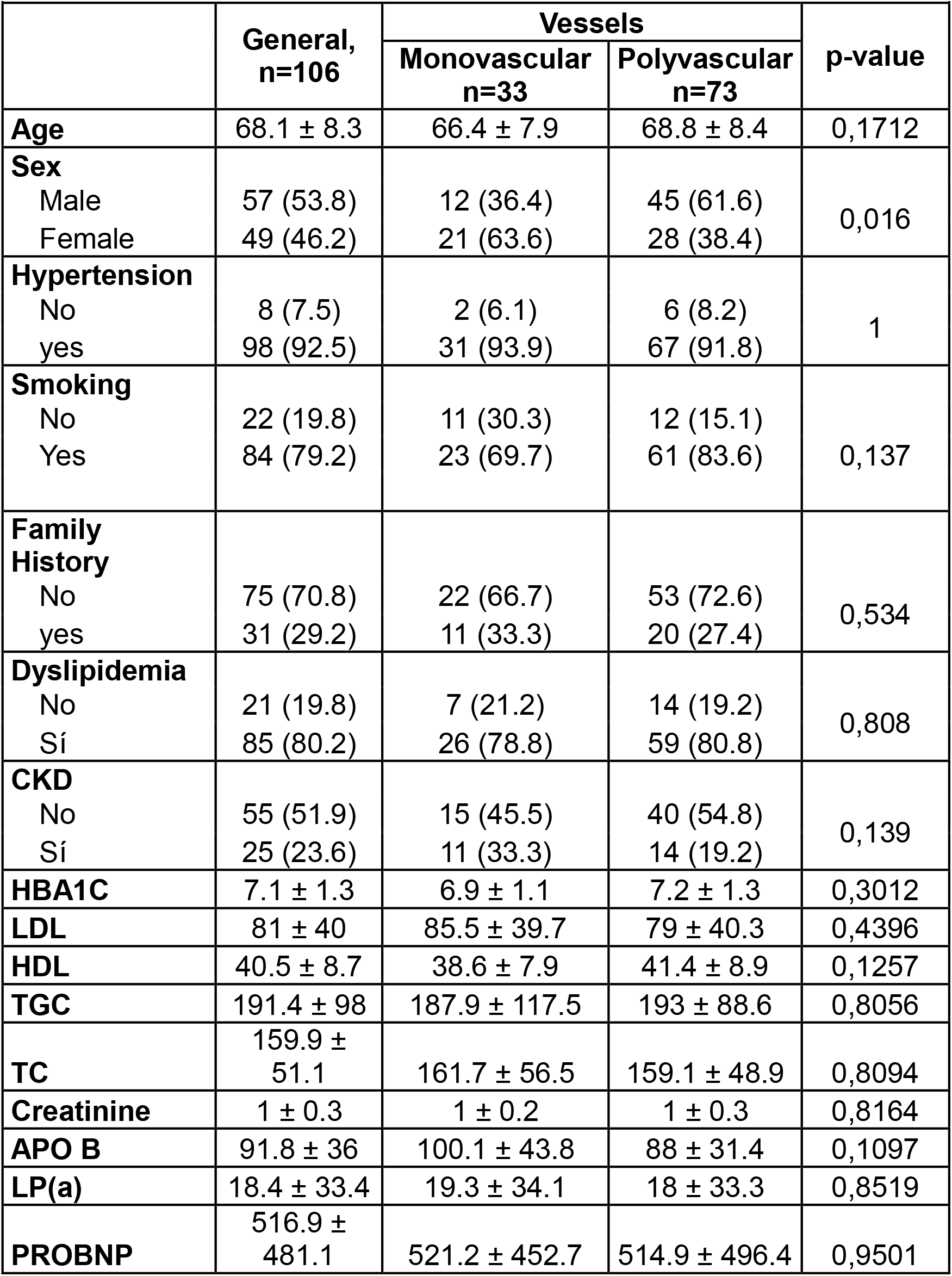
Characteristics of patients according to coronary disease complexity.

|  | General,<br>n=106 | Vessels |  | p-value |
| --- | --- | --- | --- | --- |
|  |  | Monovascular<br>n=33 | Polyvascular<br>n=73 |  |
| **Age** | 68.1 ± 8.3 | 66.4 ± 7.9 | 68.8 ± 8.4 | 0,1712 |
| **Sex** |  |  |  |  |
| Male | 57 (53.8) | 12 (36.4) | 45 (61.6) | 0,016 |
| Female | 49 (46.2) | 21 (63.6) | 28 (38.4) |  |
| **Hypertension** |  |  |  |  |
| No | 8 (7.5) | 2 (6.1) | 6 (8.2) | 1 |
| yes | 98 (92.5) | 31 (93.9) | 67 (91.8) |  |
| **Smoking** |  |  |  |  |
| No | 22 (19.8) | 11 (30.3) | 12 (15.1) | 0,137 |
| Yes | 84 (79.2) | 23 (69.7) | 61 (83.6) |  |
| **Family History** |  |  |  |  |
| No | 75 (70.8) | 22 (66.7) | 53 (72.6) | 0,534 |
| yes | 31 (29.2) | 11 (33.3) | 20 (27.4) |  |
| **Dyslipidemia** |  |  |  |  |
| No | 21 (19.8) | 7 (21.2) | 14 (19.2) | 0,808 |
| Sí | 85 (80.2) | 26 (78.8) | 59 (80.8) |  |
| **CKD** |  |  |  |  |
| No | 55 (51.9) | 15 (45.5) | 40 (54.8) | 0,139 |
| Sí | 25 (23.6) | 11 (33.3) | 14 (19.2) |  |
| **HBA1C** | 7.1 ± 1.3 | 6.9 ± 1.1 | 7.2 ± 1.3 | 0,3012 |
| **LDL** | 81 ± 40 | 85.5 ± 39.7 | 79 ± 40.3 | 0,4396 |
| **HDL** | 40.5 ± 8.7 | 38.6 ± 7.9 | 41.4 ± 8.9 | 0,1257 |
| **TGC** | 191.4 ± 98 | 187.9 ± 117.5 | 193 ± 88.6 | 0,8056 |
| **TC** | 159.9 ± 51.1 | 161.7 ± 56.5 | 159.1 ± 48.9 | 0,8094 |
| **Creatinine** | 1 ± 0.3 | 1 ± 0.2 | 1 ± 0.3 | 0,8164 |
| **APO B** | 91.8 ± 36 | 100.1 ± 43.8 | 88 ± 31.4 | 0,1097 |
| **LP(a)** | 18.4 ± 33.4 | 19.3 ± 34.1 | 18 ± 33.3 | 0,8519 |
| **PROBNP** | 516.9 ± 481.1 | 521.2 ± 452.7 | 514.9 ± 496.4 | 0,9501 |

CKD indicates chronic kidney disease, TGC triglycerides, CT total cholesterol, APOB Apolipoprotein B, Lp(a) Lipoprotein (a)

### Genetic Profile

Ninety-nine percent of the patients had three or more homozygous variants among the 11 SNPs evaluated, and 94% had more than 7 simultaneous homozygous variants. The highest frequency of homozygosity was found in the APOE gene (rs4420638) at 91.51%, ACE2 (rs2074192) at 75.48%, and CADHERIN13 (rs8055236) at 74.52%, while the lowest frequency was in the HNF1A gene (rs2259816) at 45.28%. No single genetic variation demonstrated a statistically significant association with more complex coronary artery disease. (Table 2)

**Table 2.** Variation and Nucleotide.

| **Table 2. Variation and Nucleotide** |  |  |  |
| --- | --- | --- | --- |
| **GENE/SNP** | **FREQUENCY** | **PERCENTAGE** | **CUMULATIVE** |
| **PHACTR1**<br>**RS12526453** |  |  |  |
| Heterozygous C/G | 41 | 38,68% | 38,68% |
| Homozygous C/C | 53 | 50,00% | 88,68% |
| Homozygous G/G | 12 | 11,32% | 100,00% |
| Total | 106 | 100,00% | 100,00% |
| **CDKN2A/B**<br>**RS1412830** |  |  |  |
| Heterozygous C/T | 26 | 24,53% | 24,53% |
| Homozygous C/C | 77 | 72,64% | 97,17% |
| Homozygous T/T | 3 | 2,83% | 100,00% |
| Total | 106 | 100,00% | 100,00% |
| **ACE2**<br>**RS2074192** |  |  |  |
| Heterozygous C/T | 26 | 24,53% | 24,53% |
| Homozygous C/C | 52 | 49,06% | 73,58% |
| Homozygous T/T | 28 | 26,42% | 100,00% |
| Total | 106 | 100,00% | 100,00% |
| **HNF1A**<br>**RS2259816** |  |  |  |
| Heterozygous G/T | 58 | 54,72% | 54,72% |
| Homozygous G/G | 33 | 31,13% | 85,85% |
| Homozygous T/T | 15 | 14,15% | 100,00% |
| Total | 106 | 100,00% | 100,00% |
| **CDKN2B**<br>**RS2383206** |  |  |  |
| Heterozygous A/G | 49 | 46,23% | 46,23% |
| Homozygous A/A | 15 | 14,15% | 60,38% |
| Homozygous G/G | 42 | 39,62% | 100,00% |
| Total | 106 | 100,00% | 100,00% |
| **APOE**<br>**RS4420638** |  |  |  |
| Heterozygous A/G | 9 | 8,49% | 8,49% |
| Homozygous A/A | 97 | 91,51% | 100,00% |
| Total | 106 | 100,00% | 100,00% |
| **CDKN2A/B**<br>**RS4977574** |  |  |  |
| Heterozygous A/G | 50 | 47,17% | 47,17% |
| Homozygous A/A | 23 | 21,70% | 68,87% |
| Homozygous G/G | 33 | 31,13% | 100,00% |
| Total | 106 | 100,00% | 100,00% |
| **CELSR2**<br>**RS646776** |  |  |  |
| Heterozygous C/T | 30 | 28,30% | 28,30% |
| Homozygous C/C | 2 | 1,89% | 30,19% |
| Homozygous T/T | 74 | 69,81% | 100,00% |
| Total | 106 | 100,00% | 100,00% |
| **IRS1**<br>**RS7578326** |  |  |  |
| Heterozygous A/G | 48 | 45,28% | 45,28% |
| Homozygous A/A | 50 | 47,17% | 92,45% |
| Homozygous G/G | 8 | 7,55% | 100,00% |
| Total | 106 | 100,00% | 100,00% |
| **TCF7L2**<br>**RS7903146** |  |  |  |
| Heterozygous 1/2 | 1 | 0,94% | 0,94% |
| Heterozygous C/T | 33 | 31,13% | 32,08% |
| Homozygous 1/1 | 1 | 0,94% | 33,02% |
| Homozygous C/C | 62 | 58,49% | 91,51% |
| Homozygous T/T | 9 | 8,49% | 100,00% |
| Total | 106 | 100,00% | 100,00% |
| **CADHERIN13**<br>**RS8055236** |  |  |  |
| Heterozygous G/T | 27 | 25,47% | 25,47% |
| Homozygous G/G | 78 | 73,58% | 99,06% |
| Homozygous T/T | 1 | 0,94% | 100,00% |
| Total | 106 | 100,00% | 100,00% |

Miscellaneous genetic risk profiles were established to find a plausible association with more complex coronary artery disease based on the mutation frequencies in patients with polyvascular disease, starting from the four most frequent homozygous alleles, followed by 5, 6, and 7 more frequent alleles. No statistically significant differences were found between these risk groups.

Using a Bonferroni-corrected significance threshold of 0.008 a new polygenic risk profile (PRP-5) was created according to the odds ratios (OR) previously published in the literature. A group of 5 SNPs (rs1412830-CDKN2A/B, rs2074192-ACE2, rs4420638-APOE, rs646776-CELSR2, rs7903146-TCF7L2) was identified with an OR of 5.2 and a statistically significant association (p = 0.007) for patients with polyvascular coronary artery disease (PRP-5).

Regarding the clinical characteristics of the patients about their genetic risk profile and this PRP-5, there were no differences in any variable that could infer the severity of coronary artery disease; overall, they had homogeneity in metabolic control of type 2 diabetes (T2DM), APOB, and LDL, as well as a similar burden of cardiovascular risk factors.

No statistically significant differences were found when evaluating other conditions of interest, such as the duration of T2DM, family history of cardiovascular disease, and levels of LP(a).

## Discussion

This study documented a significant group of 5 alleles with an odds ratio (OR) of 5.2, for more complex coronary artery disease in patients with T2DM independently of clinical risk factors. Understanding that the risk of recurrence and mortality in these patients is among the highest,(17,20,21) Their disease burden tends to be more diffuse, involving multiple vessels, and also compromising smaller caliber arteries. When compared to others, these patients tend to require more coronary interventions.(19,22,23)

In this study, similar to the work by Cox et al., it was demonstrated that a large number of alleles is not necessary to seek associations in such chronic diseases. They compared 13 vs. 30 alleles, and both were useful in associating with coronary artery disease(15). Verbeek et al. was another researcher who aimed to find associations with a low number of alleles; in their case, they selected 3 (rs3135506 and rs662799 in APOA5 and rs328 in LPL), which were linked to hypertriglyceridemia and were useful in discriminating patients with higher cardiovascular risk independent of metabolic status. (24) Thanassoulis et al. from the Framingham, group constructed a risk group of 13 alleles that positively associated with hard coronary artery disease (myocardial infarction, death, complexity) but not with other outcomes, which did not change when analyzing not thirteen, but sixteen alleles. When they attempted to improve prediction with 10 alleles, they found no differences. (25) No prior studies were found using polygenic risk profiles to differentiate the complexity of coronary artery disease.

A noteworthy aspect of this study is that all previous studies were based on genetic databases, while this is the first community-based cohort in hispanoamerica demonstrating not only the highly probable worldwide prevalence of alleles but also a plausible contribution to reclassifying the risk of a patient with T2DM for having more complex coronary artery disease. Another limitation of these genetic studies is the metabolic confounders that determine risk. While these alterations increase the risk of cardiovascular disease, their risk is dynamic; as metabolic control worsens, the subsequent risk increases, and their starting point for a cardiovascular event takes on another dimension. In this study, differences were found in a risk group of 5 alleles, without significant differences in family history, demographic aspects, clinical background, or metabolic factors, as there was homogeneity in levels of HbA1c, LDL, APOB, and particularly without differences in LP(a) levels, which has a significant genetic component.

One of the relevant aspects of this discussion lies in the selection of alleles. In the past, reliance was placed on “candidate genes” without yielding favorable results. After the advent of GWAS, not all authors have achieved association results. Talmud et al. analyzed a single allele on chromosome 9p21.3 without obtaining utility for predicting future cardiovascular events.(26)

The analysis of genetic risk groups for risk factors has also failed to establish specific benefits. Kathiresan et al. analyzed 9 alleles, finding an additive effect; the more present the LDL, the greater by 20 mg/dl with a decrease in HDL. Ultimately, this was associated with a 15% increased risk of coronary artery disease but did not improve discrimination beyond traditional risk factors.(27) In another study involving only women with cardiovascular disease without diabetes, Painter et al. constructed two genetic risk scores, one using 101 alleles and the other 12. By the end of the study, they were unable to discriminate risk better than traditional methods when using these scales.(28)

This study is robust as it is the first community study in Hispanoamerica demonstrating a high prevalence of homozygosity for the alleles described by genetic databases in patients with diabetes and cardiovascular disease, identifying five that could differentiate patients with more complex coronary artery disease. However, it has limitations in its scope; to consider a change in clinical practice, a temporal study is needed to evaluate this risk allele group in a clinical setting.

There is a high prevalence of homozygosity for risk alleles for cardiovascular disease in patients with type 2 diabetes. Five alleles were identified (rs1412830-CDKN2A/B, rs2074192-ACE2, rs4420638-APOE, rs646776-CELSR2, and rs7903146-TCF7L2) that may be associated with more complex coronary artery disease. These findings suggest a new hypothesis for a future case-control study.

## Data Availability

All the data referred to the manuscript is available

